# Health-related quality of life of adult COVID-19 patients following one-month illness experience since diagnosis: findings of a cross-sectional study in Bangladesh

**DOI:** 10.1101/2021.02.19.21252073

**Authors:** Md. Ziaul Islam, Baizid Khoorshid Riaz, Syeda Sumaiya Efa, Sharmin Farjana, Fahad Mahmood

**Author notes:** **Corresponding author** e-mail address (Md. Z. Islam).

## Abstract

**Background:** The pandemic coronavirus disease 2019 (COVID-19) stances an incredible impact on the quality of life of the patients. The disease not only denigrates the physical health of the patients but also affects their mental health. This cross-sectional study aimed to assess the health-related quality of life (HRQOL) of patients.

**Methods:** The study was conducted at the National Institute of Preventive and Social Medicine (NIPSOM), Dhaka, Bangladesh during the period from June to November 2020. The study enrolled 1204 adult (>18 years) COVID-19 patients diagnosed by real-time reverse transcriptase-polymerase chain reaction (RT-PCR) assay and completed the one-month duration of illness. The patients were interviewed with the CDC HRQOL-14 questionnaire to assess their HRQOL. Data were collected by telephone-interview and reviewing medical records using a semi-structured questionnaire and checklist respectively. Informed consent was obtained from each patient before data collection.

**Results:** The majority of the COVID-19 patients were males (72.3%), urban residents (50.2%), and diverse service holders (49.6%). More than one-third (35.5%) of patients had comorbidity including hypertension (55.6%), diabetes mellitus (55.6%), ischaemic heart disease (16.4%), chronic lung (12.4%), kidney (2.8%), and liver (4.2%) diseases. The mean±SD duration of physical illness was 9.83(±7.09) days, and it was 7.97(±8.12) days for mental illness. During the one-month disease course, the general health condition was excellent/very good/good in 70.1% of the patients while it was fair/poor in 29.8% of the patients. Older age, sex, and marital status were significantly associated with at least one dimension of HRQOL. Patients having symptoms of COVID-19 and comorbidity had significantly poorer HRQOL.

**Conclusion:** COVID-19 pretenses a significant impact on the HRQOL of the patients including physical and mental illness during the clinical course. Our findings suggest more pragmatic preventive, promotive, and curative measures considering illness experiences of the COVID-19 patients to restore their quality of life.

**Highlights:** Since COVID-19 was identified first in china in 2019, it has been transmitted globally and caused a significant impact on human health. A few studies have been carried out on HRQOL of COVID-19 patients and struggled with an accurate estimation of the severity of their physical and mental illness. Most of the studies recognized the poor quality of life of COVID-19 patients after the one-month disease course. Our study provides new insights on the HRQOL of the COVID-19 patients using the CDC HRQOL-14 questionnaire. We measured the HRQOL following one-month illness experience of the patients using three modules: the healthy days’ core; the activity limitations; and the healthy days’ symptoms. The study adds information regarding general health conditions including both the physical and mental health of COVID-19 patients. The study also complements information regarding the activity limitations of the patients. The study findings could contribute to designing an efficient clinical algorithm to alleviate the illness sufferings of the COVID-19 patients using a more pragmatic approach. The study conserves decisive policy implications to concoct effective interventions for improving the HRQOL of COVID-19 patients in the country and elsewhere in other countries world-wide.

## Introduction

Coronavirus is a newly emergent severe acute respiratory syndrome coronavirus 2 (SARS-CoV-2) virus, which presented as an outbreak of pneumonia of unknown cause [1]. The virus causes coronavirus disease 2019 (COVID-19), which was declared as a global emergency on 30 January 2020 and as a pandemic on 11 March 2020 by World Health Organization [1]. Several extreme public health measures like home confinement, the lockdown of cities, limited human mobilities, an extension of national holidays, and closure of academic institutions, etc. are adopted worldwide to prevent transmission. Diverse medical measures including isolation, quarantine, and hospitalisation are also espoused to minimize the health effects of COVID-19 patients. All these measures pose a negative impact on patients’ daily living including social participation and life satisfaction [2]. Concerning all these public health interventions, patient’s physical activity decreases and impairs the psychological health of the COVID-19 patients [3].

Bangladesh faces multidimensional challenges with the pandemic of novel corona virus-2 (n-Cov-2). The infection rate remained low until the end of March but saw a steep rise in April in the country. Up to 15th November 2020, total cases of COVID-19 were more than forty thousand, and total deaths were more than six thousand [4]. However, concerns have been raised because the insufficiency of testing assays may be leaving many cases undetected in the country. Being a lower-middle-income and densely populated country, Bangladesh is struggling to cope with this pandemic situation. Apart from this, the country faces significant challenges in combating COVID-19 as it also houses a million homeless and refugees in sprawling slums and refugee camps that are conducive to the spread of pandemics [5].

An infectious disease passes through a clinical course, impairs the health of the host, and poses various physical and psychological disabilities. The subjective feeling by patients of the multifaceted effect of a disease is known as Health-related quality of life (HRQOL) [6]. It is a multi-dimensional concept for examining the impact of health status on quality of life. To assess and measure HRQOL, the Centers for Disease Control and Prevention (CDC) developed a set of four “core” questions (CDC HRQOL-4). Those four items together provide a comprehensive assessment of the burden of preventable diseases, injuries, and disabilities [7].

Individuals’ physical performance and immunological stability are correlated with psychological states. Studies also reported that poor body immunity impairs the physical and mental health of the patients. The elevated psychological problems and decreased quality of life across nations and occupations are reflecting the forthcoming worst mental as well as physical health situations to the victims of infectious diseases [8]. As a highly infectious disease, COVID-19 affects the health and quality of life of patients who are likely to have a lower HRQOL [9].

Globally, experts identified the need to pay specific attention to the COVID-19 patients and people at risk of further deterioration of physical health and mental distress that may need tailored interventions. Special attention must be paid to the patients who have preexisting psychiatric conditions, pregnant women, elderly, immunocompromised people, persons in detention, international migrant workers, and international students. Therefore, along with efforts at various levels to prevent the spread of the disease, an intervention must be a part of the public health response to restore the physical and mental health of the COVID-19 patients [10].

To date, no study has investigated how severe the impact of COVID-19 is on the quality of life of the patients in the context of Bangladesh. The current study was the pioneering invention to assess the HRQOL of the COVID-19 patients using the measuring tool of CDC. The present study intended to assess the HRQOL of adult COVID-19 patients considering both physical and mental illness in the local context of Bangladesh.

## Materials and Methods

### Study setting and design

We conducted this cross-sectional study at the National Institute of Preventive and Social Medicine (NIPSOM), Dhaka, Bangladesh during the period from June to November 2020. NIPSOM is the apex public health institute in the country holding the central laboratory designated for COVID-19 diagnosis and approved by the Ministry of Health and Family Warfare of the government of Bangladesh. The study population included all adult COVID-19 patients diagnosed by real-time reverse transcriptase-polymerase chain reaction (RT-PCR) assay at the central laboratory of NIPSOM. We enrolled the patients of both sexes, aged >18 years and who completed the one-month duration of illness after diagnosis. The patients who had no contact number (telephone/cell phone); who didn’t respond to a phone call on three separate occasions; who were unwilling to participate; and who had incomplete interviews were excluded.

### Sample size and sampling

We retrieved the COVID-19 patients using the records of the central laboratory of NIPSOM. All the COVID-19 patients (1342) diagnosed by the laboratory during the data collection period (1st to 31st July 2020) formed the sampling frame, and each of them was a sampling unit. The study enrolled the patients having criteria such as; (i) completion of the one-month duration of illness; (ii) adults aged >18 years; (iii) capable of giving informed consent, and (iv) absence of severe illness. Of all, a total 1204 of COVID-19 patients were enrolled in the study using the purposive sampling technique.

### Data collection

We used a semi-structured questionnaire and a checklist for data collection through telephone interviews and records review. The questionnaire comprised variables related to baseline characteristics, health-related quality of life, symptoms, and commodities associated with COVID-19. A checklist was used to collect laboratory data of the CIVID-19 patients by reviewing medical records obtained from the central laboratory of NIPSOM. Both data collection instruments were pretested on the COVID-19 patients diagnosed in June 2020 and accordingly, necessary corrections and modifications were performed for its finalization. To ensure the validity of data, we recorded each telephone interview by a digital recorder along with filling up the hard copy of the questionnaire. Participation of the COVID-19 patients was voluntary and informed consent was obtained from each participant before data collection. Before taking consent, each participant was informed about the objectives and procedure of the study along with the risks and benefits of participation in the study. Measures were taken to ensure the quality, consistency, and relevancy of data.

### CDC HRQOL-14

The CDC HRQOL-14 questionnaire was used to measure the HRQOL of the COVID-19 patients. The questionnaire consists of 3 modules: i) healthy days’ core module comprising of 4 questions; ii) activity limitations module comprising of 5 questions, and iii) healthy days’ symptoms module comprising of 5 questions [11].

### Statistical analysis

Data were analysed using SPSS STATISTICS (Version 25.0, IBM Statistical Product and Service Solutions, Armonk, NY, USA). The normality of the variables was tested with the Shapiro Wilk test/Kolmogorov Smirnov tests of Normality. Continuous data were portrayed in the form of mean and standard deviation. Categorical data were depicted as frequency and percentages. Descriptive statistics estimated mean, standard deviation, and frequency while inferential statistics included chi-square test, independent sample t-test, and F-test. We used the chi-square test to compare the significant difference between two categorical variables. The t-test was used to find a significant difference of means of two quantitative variables and the F-test was used to find a significant difference of means of three or more quantitative variables. Significant F-test results were further analyzed by Post-Hoc test, and significant categorical data were analyzed by logistic regression analysis. A p-value <0.05 was considered significant with 95% confidence interval (CI). All the statistical tests were two-sided and were performed at a significance level of α= 0.05.

### Ethical considerations

We maintained all kinds of ethical issues in different stages of the study following the guidelines of Helsinki declarations for conducting any study using human subjects. Informed consent was obtained from the participants before data collection. During data collection, the confidentiality of data and privacy of the participants were maintained strictly. Participation of the COVID-19 patients was voluntary, and they had the freedom to withdraw their consent at any stage of the study. We used the collected data anonymously only for the current study.

## Results

Out of 1342 COVID-19 patients, 1204 (89.7%) participated in the study. Followed by 5.1% of the patients were with a wrong contact number, 2.1% didn’t attend the phone calls, 1.9% were unwilling to participate, and 1.2% had an incomplete interview (Fig. 1).

**Fig. 1.**
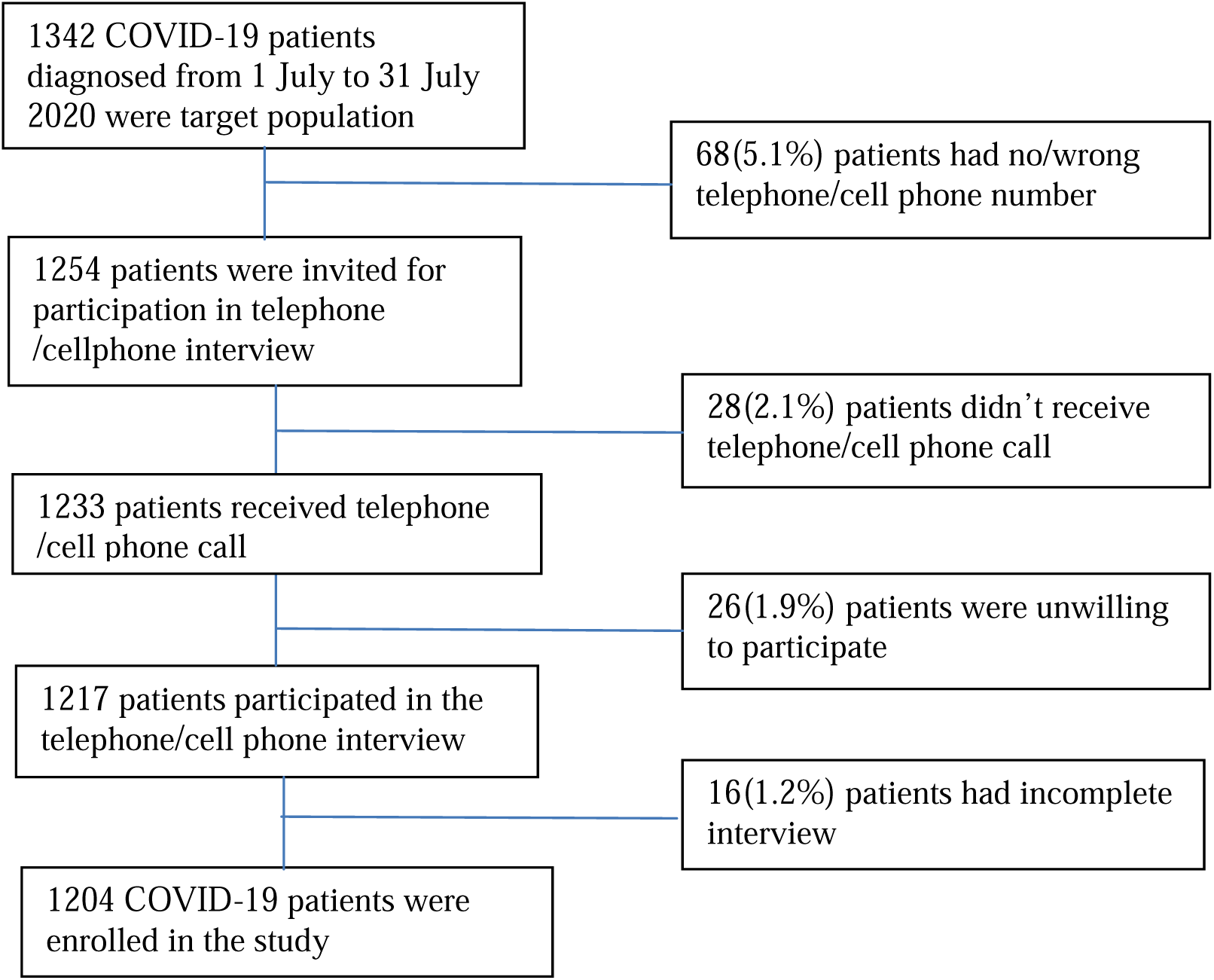
Flow chart of the study participants (COVID-19 patients)

The majority (49.3%) of the patients were in the age group 30–49 years followed by 20.8% belonged to the age group 20–29 years, and the mean (SD) age of the patients was 41.63 (±13.53) years. Around three-fourth (72.3%) of patients were males, and the rest 27.7% were females. Of all, 82.8% were currently married, 25.1% completed their graduate-level education, and 49.6% were service holders. Almost equal numbers of patients were from urban and rural areas (49.8% vs. 50.2%). More than two-thirds (67.5%) were from a nuclear family. The majority (48.3%) of the patients had monthly family income between TK.21000 and TK.50000, and their average monthly family income was TK.32229.64±25797.41 (Table 1).

**Table 1.**
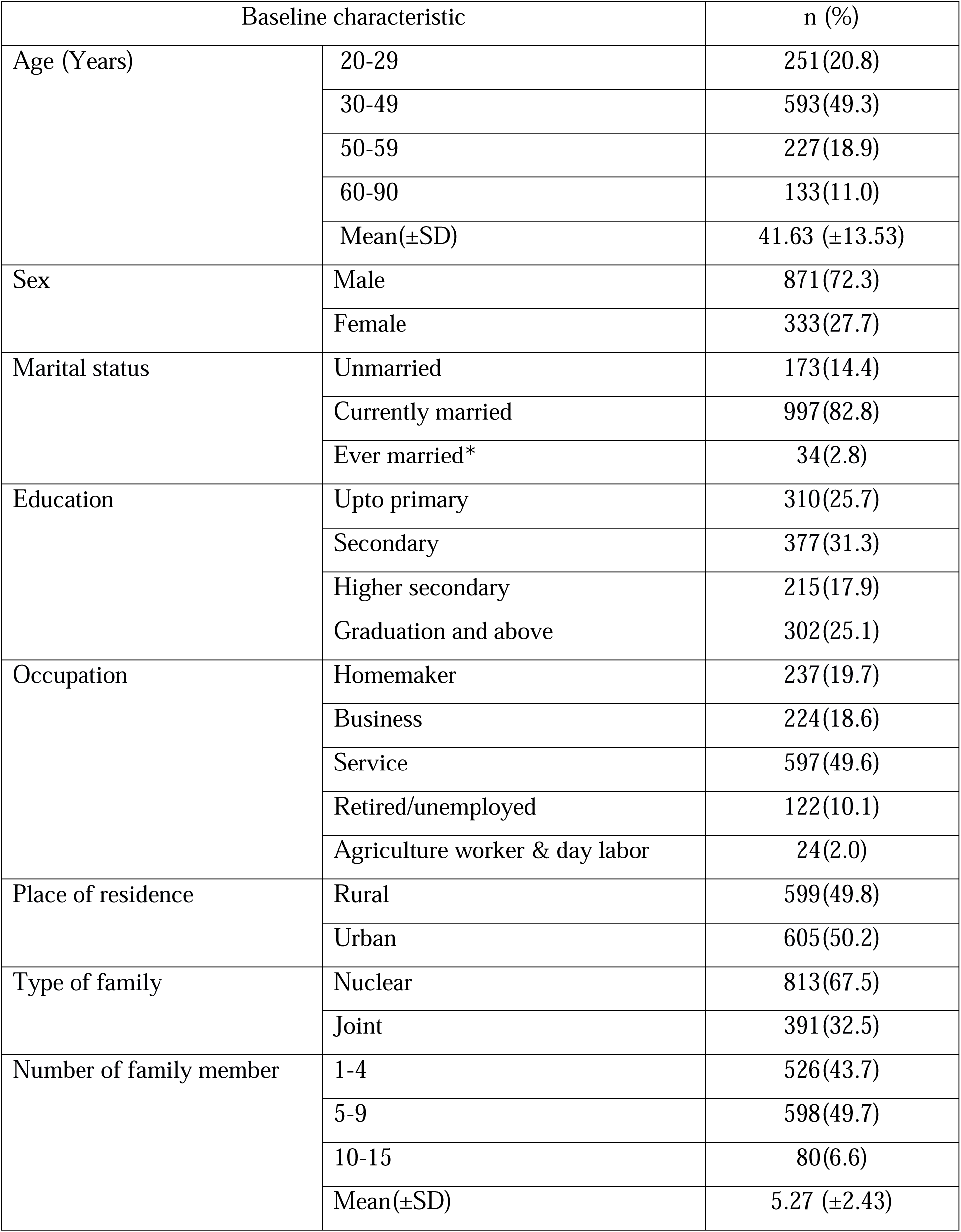
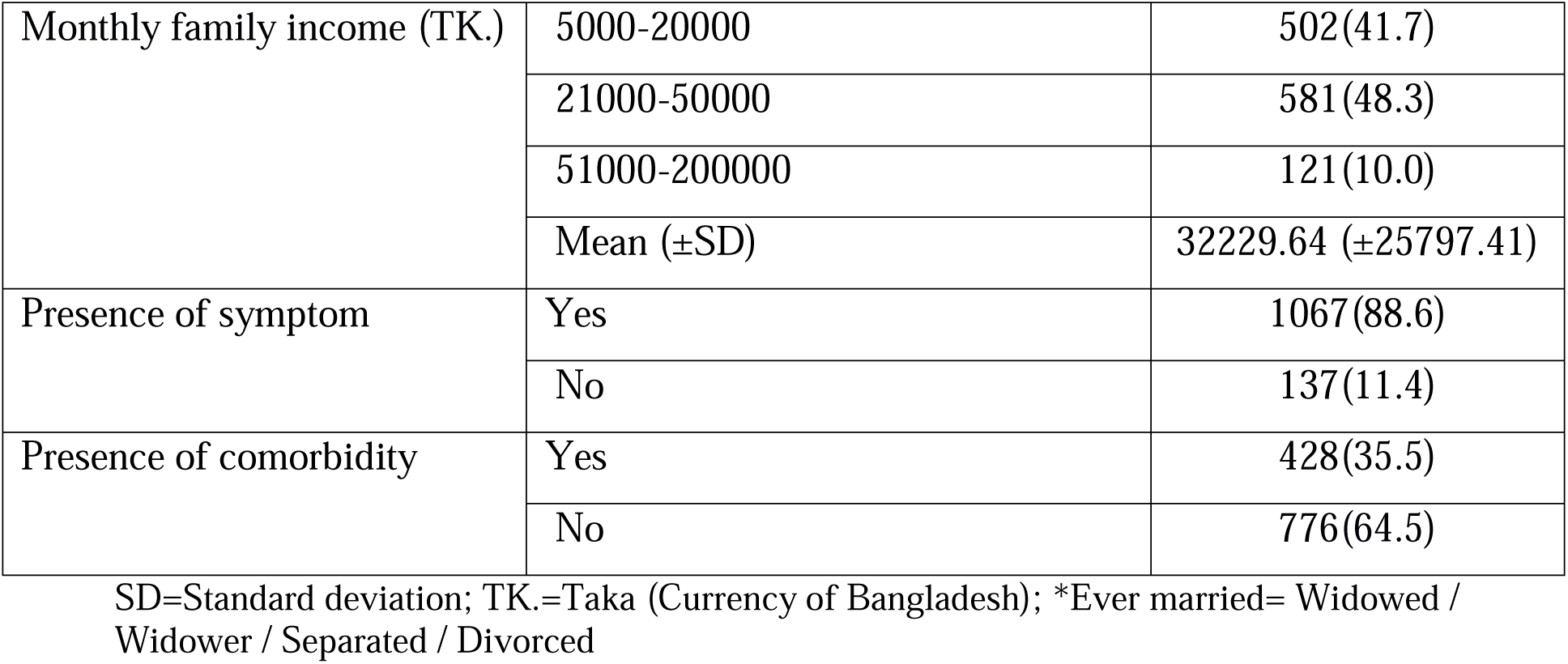
Distribution of the COVID-19 patients by baseline characteristics.

Concerning symptoms of COVID-19, the majority (86.0%) of the patients had a fever. Other symptoms included cough (59.0%), sore throat (41.0%), anosmia (38.2%), shortness of breath (26.0%), and diarrhea (16.1%) (Fig. 2). Out of all 1204 patients, 35.5% COVID-19 patients had comorbidity. Regarding the type of comorbidity, 55.6% of the patients had hypertension, and another 55.6% had diabetes mellitus. Another 16.4% of the patients had ischemic heart disease, and 12.4% had lung disease. Only 4.2% of the patients had chronic liver disease, and 2.8% had chronic kidney disease (Fig. 3).

**Fig. 2.**
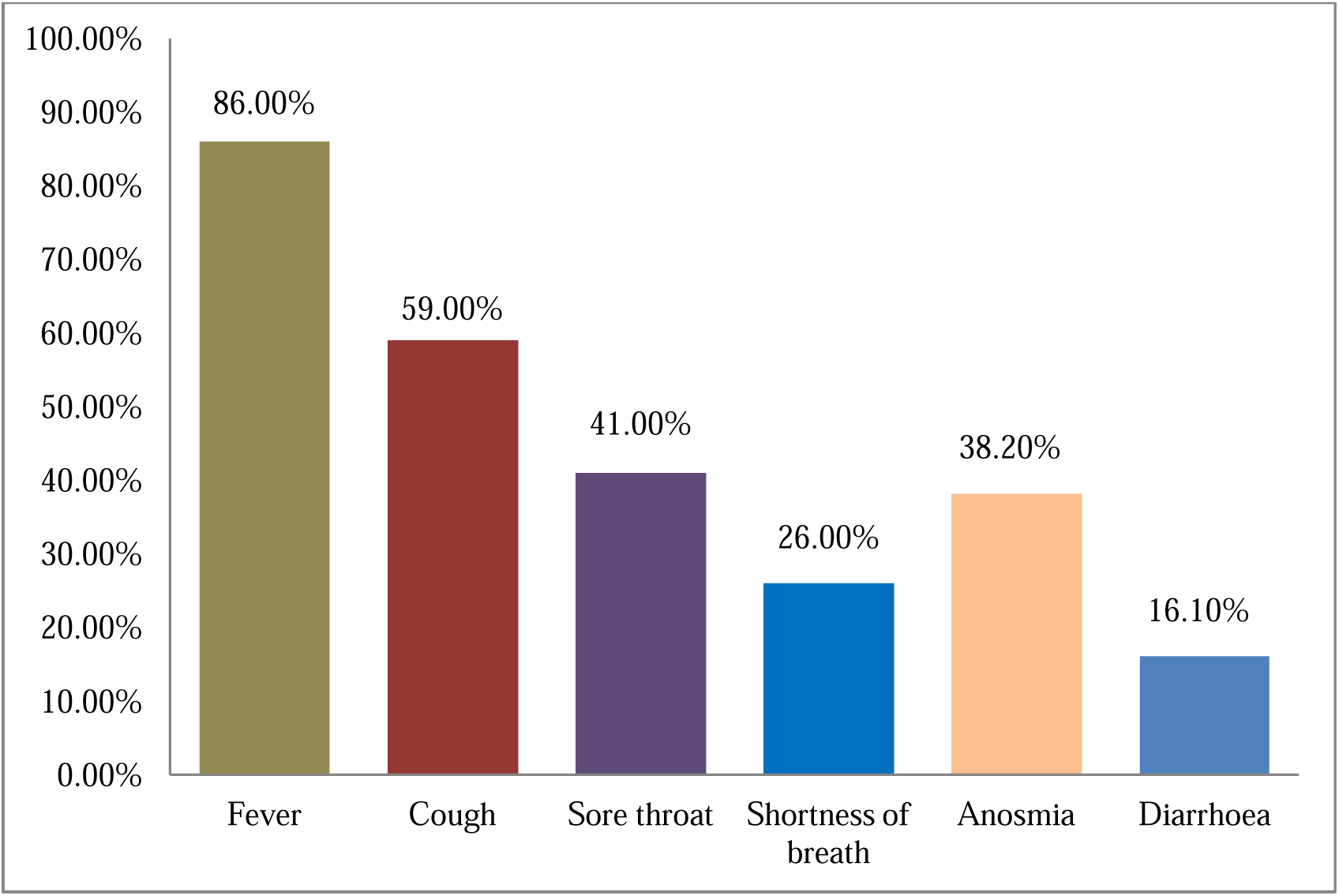
Distribution of the symptoms of the COVID-19 patients.

**Fig. 3.**
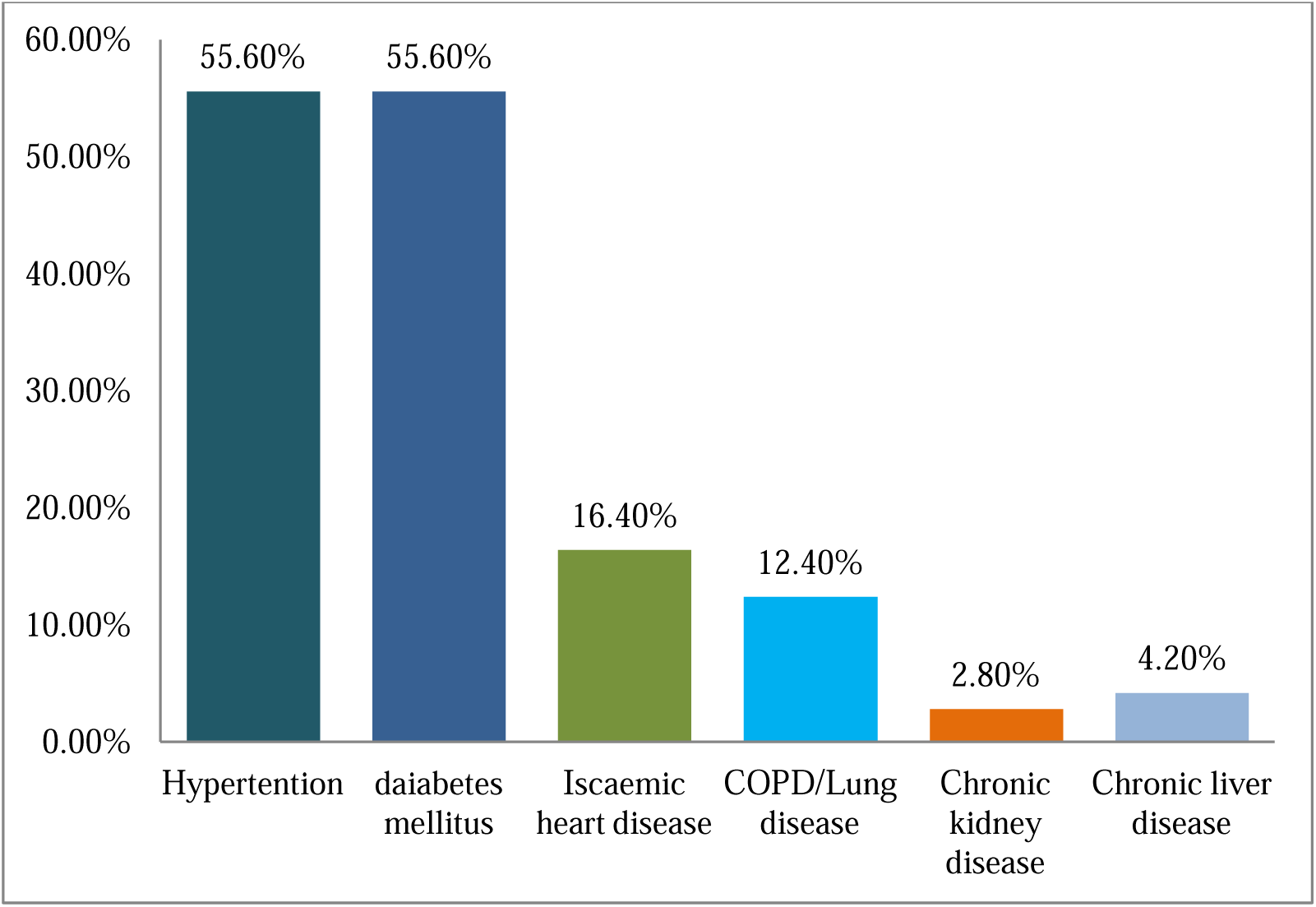
Distribution of the COVID-19 patients by comorbid conditions (n=427)

The general health condition was excellent/very good/good in 70.1% of the patients, and it was fair/poor in 29.8% of the patients. The majority (87.0%) of the COVID-19 patients needed help for personal care, and 47.8% required help for routine needs. The mean (±SD) duration of physical illness of the patients was 9.83(±7.09) days, and that of mental illness was 7.97(±8.12) days. The mean (±SD) duration usual activity limitation was 7.04(±5.53) days while the mean (±SD) duration of health-related limited activity was 11.39(±9.29) days. The study also estimated the mean (±SD) duration of hard to do usual activities due to pain (3.88±7.01 days), feeling sad or depressed (7.04±8.07) days), feeling worried (6.77±8.03 days), not getting enough rest (4.38±6.79 days), and feeling healthy (14.83±9.49) days (Table 2).

**Table 2.** Distribution of different items of health-realted quality of life of the COVID-19 patients (based on CDC HRQOL-14)

| **Items of healthy days core module** |  |  |
| --- | --- | --- |
| Patient's general health condition, n (%) | Excellent/very good/good | 845(70.1) |
|  | fair/poor | 359(29.8) |
| Duration of physical illness (Mean±SD days) |  | 9.83(±7.09) |
| Duration of mental illness (Mean±SD days) |  | 7.97(±8.12) |
| Duration of usual activity limitation (Mean±SD days) |  | 7.04(±5.53) |
| **Items of activity limitations module** |  |  |
| Duration of health related limited activity (Mean±SD days) |  | 11.39(±9.29) |
| Need help for personal care, n (%) | Yes | 1047(87.0) |
|  | No | 157(13.0) |
| Need help for routine needs, n (%) | Yes | 576(47.8) |
|  | No | 628(52.2) |
| **Items of healthy days symptoms module** |  |  |
| Duration of hard to do usual activities due to pain (Mean±SD days) |  | 3.88(±7.01) |
| Duration of feeling sad or depressed (Mean±SD days) |  | 7.04(±8.07) |
| Duration of feeling worried (Mean±SD days) |  | 6.77(±8.03) |
| Duration of not getting enough rest (Mean±SD days) |  | 4.38(±6.79) |
| Duration of feeling healthy (Mean±SD days) |  | 14.83(±9.49) |

SD= Standard Deviation; n= Number; %= Percentage

The risk of fair/poor general health condition was significantly associated with the presence of symptoms (OR=33.22, CI=8.17-135.1) and having comorbidity (OR=1.573, CI=1.18-2.10). Patients’ need for help for routine needs was also significantly associated with having symptom (OR=2.730, CI=1.82-4.09) and having comorbidity (OR=1.536, CI=1.18-2.0) (Table 3).

**Table 3.**
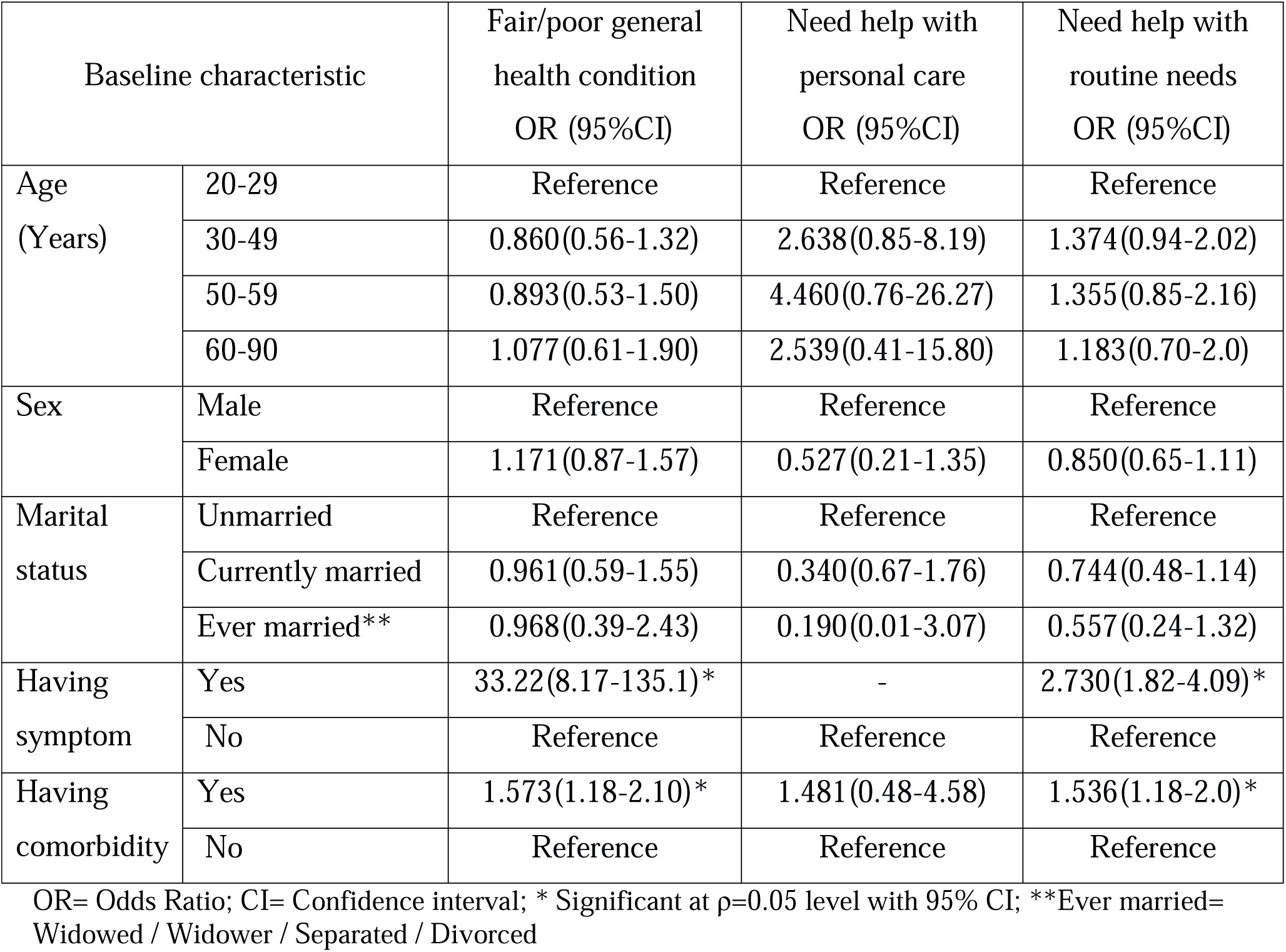
Logistic regression analysis of items of HRQOL by baseline and clinical characteristics of the the COVID-19 patients.

| Baseline characteristic |  | Fair/poor general health condition<br>OR (95%CI) | Need help with personal care<br>OR (95%CI) | Need help with routine needs<br>OR (95%CI) |
| --- | --- | --- | --- | --- |
| Age (Years) | 20-29 | Reference | Reference | Reference |
|  | 30-49 | 0.860(0.56-1.32) | 2.638(0.85-8.19) | 1.374(0.94-2.02) |
|  | 50-59 | 0.893(0.53-1.50) | 4.460(0.76-26.27) | 1.355(0.85-2.16) |
|  | 60-90 | 1.077(0.61-1.90) | 2.539(0.41-15.80) | 1.183(0.70-2.0) |
| Sex | Male | Reference | Reference | Reference |
|  | Female | 1.171(0.87-1.57) | 0.527(0.21-1.35) | 0.850(0.65-1.11) |
| Marital status | Unmarried | Reference | Reference | Reference |
|  | Currently married | 0.961(0.59-1.55) | 0.340(0.67-1.76) | 0.744(0.48-1.14) |
|  | Ever married** | 0.968(0.39-2.43) | 0.190(0.01-3.07) | 0.557(0.24-1.32) |
| Having symptom | Yes | 33.22(8.17-135.1)* | - | 2.730(1.82-4.09)* |
|  | No | Reference | Reference | Reference |
| Having comorbidity | Yes | 1.573(1.18-2.10)* | 1.481(0.48-4.58) | 1.536(1.18-2.0)* |
|  | No | Reference | Reference | Reference |

OR= Odds Ratio; CI= Confidence interval; \* Significant at $p=0.05$ level with 95% CI; \*\*Ever married= Widowed / Widower / Separated / Divorced

The mean duration of “not good” physical health was significantly (p<0.05) higher in the age group 60-90 years (11.08 days). The mean duration of “not good” physical health was significantly (p<0.05) higher among patients having symptoms (11.09 days) and having comorbidity (11.07 days). The mean duration of “not good” mental health was significantly (p<0.05) higher in females (8.73 days), ever married patients (10.44 days), and patients having symptoms (8.67 days) and having comorbidity (8.78 days). The mean duration of ‘usual activity limitation’ was significantly (p<0.05) higher in the age group 50-59 years (7.88 days), patients having symptoms (7.4 days), and comorbidity (7.88 days). The mean duration of ‘health-related limited activity’ was significantly (p<0.05) higher in the age group 50-59 years (7.88 days). The mean duration of ‘health-related limited activity’ was also significantly (p<0.05) higher in the patients having symptoms (7.94 days) and comorbidity (7.88 days) (Table 4).

**Table 4.** Mean±SD duration of selected items of HRQOL by baseline and clinical attributes of the COVID-9 patients.

| Attributes |  | Duration of “Not good” physical health (Mean±SD days) | Duration of “Not good” mental health (Mean±SD days) | Duration of usual activity limitation (Mean±SD days) | Duration of health –related limited activity (Mean±SD days) |
| --- | --- | --- | --- | --- | --- |
| Age (Years) | 20-29 | 8.65(6.74)<sup>a</sup> | 7.50(7.58)<sup>a</sup> | 11.09(9.56)<sup>a</sup> | 6.12(5.0)<sup>a</sup> |
|  | 30-49 | 9.71(7.14) | 7.89(8.09) | 11.80(9.33) | 7.06(5.64) |
|  | 50-59 | 10.71(6.84)* | 7.95(8.22) | 11.34(8.98) | 7.88(5.85)* |
|  | 60-90 | 11.08(7.65)* | 9.24(8.99) | 10.17(9.06) | 7.26(5.26) |
| Sex | Male | 9.84(6.96) | 7.68(8.01)* | 11.75(9.31)* | 7.14(5.35) |
|  | Female | 9.81(7.44) | 8.73(8.38)* | 10.44(9.18)* | 6.78(5.99) |
| Marital status | Unmarried | 8.57(7.01) | 6.62(6.85)<sup>a</sup> | 11.86(9.75) | 6.34(5.41) |
|  | Currently married | 10.06(7.11) | 8.12(8.28) | 11.38(9.26) | 7.19(5.59) |
|  | Ever married** | 9.62(6.61) | 10.44(8.63)* | 9.12(7.31) | 6.09(4.25) |
| Having symptom | Yes | 11.09(6.54)* | 8.67(8.16)* | 12.23(9.08)* | 7.94(5.23)* |
|  | No | 0.0(0.0) | 2.55(5.25) | 4.82(8.20)* | 0.0(0.0) |
| Having comorbidity | Yes | 11.07(7.31)* | 8.78(8.51)* | 10.88(9.30) | 7.88(5.97)* |
|  | No | 9.15(6.88) | 7.52(7.87) | 11.66(9.28) | 6.57(5.23) |

SD= Standard deviation; <sup>a</sup> Reference category; \* t-test/Post-Hoc test, Significant at $p<0.05$ level with 95% CI; \*\*Ever married= Widowed / Widower / Separated / Divorced

The mean duration of ‘feeling pain’ was significantly associated with the presence of symptoms (4.33 days) and having comorbidity (5.28 days). The mean duration of ‘feeling sad, blue, or depressed’ was significantly (p<0.05) higher in ever married patients (10.47 days), patients having symptoms (7.58 days), and comorbidity (8.04 days). The mean duration of ‘feeling worried’ was significantly (p<0.05) associated with gender (male=6.48 days and female=7.53 days) and presence of symptom (7.26 days). The mean duration of ‘feeling worried’ was significantly (p<0.05) higher among ever married patients (10.12 days). The mean days of ‘not getting enough rest’ were significantly (p<0.05) higher in females (5.26 days), having symptoms (4.69 days) and comorbidity (5.02 days). But the mean duration of ‘feeling very healthy’ was significantly (p<0.05) lower in the age group 50-59 years (13.24 days). The mean duration offeeling very healthy’ was also significantly (p<0.05) lower in the patients having symptoms (13.51 days) and comorbidity (13.0 days) (Table 5).

**Table 5.** Mean±SD duration of selected items of HRQOL by baseline and clinical attributes of the COVID-19 patients.

| Attributes |  | Feeling pain<br>(Mean±SD<br>days) | Feeling sad,<br>blue or<br>depressed<br>(Mean±SD<br>days) | Feeling<br>worried<br>(Mean±SD<br>days) | Not getting<br>enough rest<br>(Mean±SD<br>days) | Feeling very<br>healthy<br>(Mean±SD<br>days) |
| --- | --- | --- | --- | --- | --- | --- |
| Age (Years) | 20-29 | 2.94(6.25)<sup>a</sup> | 6.39(7.59)<sup>a</sup> | 6.20(7.41)<sup>a</sup> | 4.16(6.61)<sup>a</sup> | 16.07(9.63)<sup>a</sup> |
|  | 30-49 | 3.92(6.97) | 6.98(7.90) | 6.77(7.89) | 4.66(7.13) | 15.14(9.25) |
|  | 50-59 | 4.59(7.27) | 7.20(8.30) | 6.90(8.36) | 3.83(5.86) | 13.24(9.37)* |
|  | 60-90 | 4.28(7.89) | 8.28(9.18) | 7.59(9.14) | 4.53(7.05) | 13.80(10.08) |
| Sex | Male | 3.69(6.67) | 6.86(7.95) | 6.48(7.90)* | 4.05(6.35)* | 15.03(9.43) |
|  | Female | 4.40(7.80) | 7.51(8.37) | 7.53(8.33)* | 5.26(7.76)* | 14.03(9.62) |
| Marital<br>status | Unmarried | 2.83(6.38) | 5.92(7.04)<sup>a</sup> | 5.57(6.80)<sup>a</sup> | 3.64(6.22) | 16.37(9.60) |
|  | Currently<br>married** | 4.08(7.13) | 7.12(8.18) | 6.86(8.16) | 4.46(6.83) | 14.64(9.48) |
|  | Ever married | 3.38(5.83) | 10.47(8.85)* | 10.12(9.06)* | 5.82(8.12) | 12.53(8.25) |
| Having<br>symptom | Yes | 4.33(7.28)* | 7.58(8.19)* | 7.26(8.20)* | 4.69(6.89)* | 13.51(8.74)* |
|  | No | 0.39(2.17) | 2.86(5.56) | 2.91(5.17) | 2.01(5.43) | 25.05(8.84) |
| Having<br>comorbidity | Yes | 5.28(8.29)* | 8.04(8.51)* | 7.36(8.34) | 5.02(7.32)* | 13.0(9.86)* |
|  | No | 3.12(6.06) | 6.49(7.77) | 6.44(7.84) | 4.03(6.45) | 15.84(9.13) |

SD= Standard deviation; <sup>a</sup>Reference category; \*t-test/Post-Hoc test, Significant at $p=0.05$ level with 95% CI; \*\*Ever married= Widowed / Widower / Separated / Divorced

## Discussion

This cross-sectional study appraised the HRQOL of adult COVID-19 patients in the local context of Bangladesh. Following the case definition of COVID-19 mentioned in the national guideline of Bangladesh, the study enrolled COVID-19 patients confirmed by the RT-PCR test [12]. The current study was the pioneering invention to assess the health-related quality of COVID-19 patients using CDC-HRQOL in the national and international platforms. The HRQOL comprised general health condition, physical illness, mental illness, and usual activity limitation following the experiences of the COVID-19 patients in the one-month clinical course of the disease since diagnosis. The study findings preserve potential policy implications to contrive expedient guidelines for effective clinical management of the patients to refurbish their quality of life and health condition. The study also conserves enormous academic and research implications to track the clinical course of the COVID-19 and its impacts on the physical and mental health of the patients. Accordingly, the study could contribute to the reform and redesign of the healthcare delivery system for the COVID-19 patients.

In our study, around three-fourth of the COVID-19 patients were males, and the rest were females. This finding indicates that males were at a higher risk of being infected by COVID-19 than females. In this regard, other studies conducted in China [13] and Vietnam [14] revealed reverse findings where females were affected more than males. It may be due to the divergence of the socio-cultural context of Bangladesh, which is different from those countries. In the existing social and cultural frame of the country, males are more involved in outdoor activities than females. So, the chance of exposure and being affected are higher in males than the females. Around half of the COVID-19 patients were young adults (age group 30-49 years) and only 11.0% of the patients were elderly (age group 60-90 years). The demographic profile of Bangladesh depicts that 40.07% of the population belongs to the age group of 25–54 years and only 6.42% belongs to the age group >60 years [1]. In this regard, it could be claimed that the age distribution of the patients is in alignment with the national age distribution.

An almost equal number of the COVID-19 patients reported from the urban (50.2%) and rural (49.8%) communities. Another study conducted by Islam MZ et al in Bangladesh [1] revealed different findings where patients were reported more from the urban communities. Though these two studies were conducted in the same country, a different result may be due to differences in the study periods. The former study was conducted earlier in comparison to the current one. At the beginning of the COVID-19 pandemic, the diagnostic facility was limited in the urban setting of Bangladesh. Moreover, rural people of the country were less aware of the COVID-19 diagnosis and personal protection. As a result, the former study found more patients reported from the urban setting than our study.

By occupation, nearly half of the patients were service holders. In this respect, the study conducted by Islam MZ et al [1] also found the majority of the patients as service holders but it was relatively lower (32.5%) than the current study. It could be justified by the reality that a remarkable segment of the people of Bangladesh are involved in diverse private and public jobs for their survival, and they have to move outside to attend their job stations. As a result, the service holders are more vulnerable to the get infected by COVID-19 than other occupational groups. It could also be mentioned that during the early period of the pandemic, people were restricted to social movement due to lockdown, shut down, quarantine, and the panic situation caused by the disease. It could be a valid reason for comparatively fewer service holders in the former study than the current study.

Among all the patients under the current study, more than four-fifth (86.0%) had various types of symptoms of COVID-19. This finding indicates that the majority of the patients were symptomatic and attended health facilities for diagnosis of the disease. It was also revealed that a sensible part (14.0%) of the patients were asymptomatic but attended the health facilities for diagnosis of the disease due to their exposure history, treatment of comorbidity, travel history, and other indications related to COVID-19 infection. The present study also portrayed that more than one-third (35.5%) of the COVID-19 patients had different comorbidities including hypertension, diabetes mellitus, ischaemic heart disease, chronic lung, kidney, and liver diseases. The former study conducted by Islam MZ et al [1] also identified nearly the same proportion (33.9%) of the COVID-19 patients having comorbidities including hypertension, coronary heart disease, diabetes mellitus, cancers, chronic lung, kidney, and kier diseases. The comorbid patients were at higher risk of worse morbidity and mortality consequences of COVID-19, and they were referred to health facilities to exclude COVID-19.

We retrieved the data of the COVID-19 patients who had completed their one-month duration of illness using their laboratory records. At the end of one month, the general health condition was excellent/very good/good in 70.1% patients while it was fair/poor in the rest 29.8% patients. Fair/poor health condition was significantly associated with having a symptom (OR=33.22, CI=8.17-135.1) and having any comorbidity (OR=1.573, CI=1.18-2.10). It could be explained by the fact that the symptoms aggravate the morbidity severity and illness feelings of the patients, and thus interfere with their general health condition.

The current study estimated the average duration of physical illness around ten days, and this duration increased proportionately with the increase in age of the patients. On the other hand, the average duration of mental illness was around eight days, and this duration was significantly higher among females and ever married patients. Arguably it could be mentioned that the females and elderly ever married patients conserve relatively less body immunity to protect the illness progression and its clinical severity. As a result, their duration of suffering lasts longer than their counterpart males and younger adults. The average duration of usual activity limitation was around seven days, and it was significantly higher in the age group 50-59 years. The presence of comorbidity poses an incremental effect on the disease progression and worsens their health condition. In the present study, patients’ need for help for routine needs was also significantly higher in the patients having a symptom (OR=2.730, CI=1.82-4.09) and comorbidity (OR=1.536, CI=1.18-2.0). The duration of health-related limited activity was also significantly higher in the age group 50-59 years and the patients having symptoms and comorbidity. All items of the healthy days’ core module (duration of physical and mental illness, usual activity limitation) were significantly higher among patients’ having a symptom and comorbidity. It is evident that several clinical manifestations of symptoms along with comorbidity deteriorate both the physical and mental health condition of the patients, which decrease their ability to performing daily usual activities.

The mean duration (in days) of feeling pain was significantly higher in the patients having symptoms and comorbidity. The mean duration of feeling sad, blue, or depressed was significantly higher in the ever married patients, and patients having symptoms and comorbidity. On the contrary, the mean duration of feeling worried was significantly higher in female patients and the patients having symptoms. The mean duration of feeling worried was significantly higher in the ever married patients while the mean duration of not getting enough rest was significantly higher in females and the patients having symptom and comorbidity. The mean duration of feeling very healthy was significantly lower in the age group 50-59 years, and the patients having symptoms and comorbidity. The findings of the present study revealed that the mean duration of symptoms was significantly higher in the females, ever married, and the patients having symptoms and comorbidity. All these findings entice special attention of the health policymakers and healthcare managers to invent prioritized medical measures for the COVID-19 patients to improve their HRQOL and general health condition.

## Conclusion

The current study concerns a limitation with telephone interview might not address some more precise information, which could be unveiled from the patients by face-to-face interview. The sample size was adequate for exploring the interactions that provide substantial evidence and directions for future instructions and public health interventions to reorganize medical management modalities for the COVID-19 patients. The study findings also recommend revising national guidelines for clinical management of COVID-19 patients based on the factors associated with HRQOL. Special attention must be paid to the determinants of physical and mental illness, and the limitation of daily activities of the COVID-19 patients.

## Data Availability

Individual participant data that underlie the results reported in this article, after de-identification (text, tables, figures, and appendices) will be available immediately following publication indefinitely, to anyone who wishes to access the data, for any purpose. Individuals who would like to access the data should contact the corresponding author.

## Author contributions

MZI had the idea for the study. MZI, BKR, SSE, SF and FM equally contributed in study design and literature review. All authors had involved in the data collection. MZI ans SSE did the data analysis. All authors interpreted the data analysis and helped in drafting the manuscript. All authors have approved the final manuscript.

## Funding statement

This research did not receive any specific grant from funding agencies in the public, commercial, or not-for-profit sectors.

## Competing interests

The authors have no competing interests.

## Ethical approval

Ethical approval (Ref. No. NIPSOM/IRB/2020/5) was obtained from the Institutional Review Board (IRB) of the National Institute of Preventive and Social Medicine (NIPSOM), Dhaka, Bangladesh.

## Acknowledgments

All authors would like to acknowledge the laboratory authority, medical technologists, and supporting staffs for their continuing and unconditional support in sharing laboratory records of the diagnosed patients. We also forward sincere appreciation to the patients and their families for their unrestricted supports in data collection through a telephone interview.

## Notes

### Competing Interest Statement

The authors have declared no competing interest.

## References

1. Islam MZ, Riaz BK, Islam AN, Khanam F, Akhter J, Choudhury R, Farhana N, Jahan NA, Uddin MJ, Efa SS. Risk factors associated with morbidity and mortality outcomes of COVID-19 patients on the 28th day of the disease course: a retrospective cohort study in Bangladesh. Epidemiology & Infection. 2020;148. Available form: https://doi.org/10.1017/S0950268820002630.

2. Qi M, Li P, Moyle W, Weeks B, Jones C. Physical activity, health-related quality of life, and stress among the Chinese adult population during the COVID-19 pandemic. International Journal of Environmental Research and Public Health. 2020 Jan;17(18):6494.

3. Suzuki Y, Maeda N, Hirado D, Shirakawa T, Urabe Y. Physical activity changes and its risk factors among community-dwelling japanese older adults during the COVID-19 epidemic: Associations with subjective well-being and health-related quality of life. International Journal of Environmental Research and Public Health. 2020 Jan;17(18):6591.

4. World Health Organization (WHO). COVID-19, Mortality and Morbidity Weekly Update (MMWU). Accessed on 16thNovember, 2020. https://www.who.int/southeastasia/outbreaks-and-emergencies/novel-coronavirus-2019.

5. Anwar S, Nasrullah M, Hosen MJ. COVID-19 and Bangladesh: Challenges and how to address them. Frontiers in Public Health. 2020;8. Available form: https://dx.doi.org/10.3389%2Ffpubh.2020.00154

6. Chen KY, Li T, Gong F, Zhang JS, Li XK. Predictors of health-related quality of life and influencing factors for COVID-19 patients, a follow-up at one month. Frontiers in Psychiatry. 2020;11:668.

7. Yin S, Njai R, Barker L, Siegel PZ, Liao Y. Summarizing health-related quality of life (HRQOL): development and testing of a one-factor model. Population health metrics. 2016 Dec 1;14(1):22.

8. Islam SD, Bodrud-Doza M, Khan RM, Haque MA, Mamun MA. Exploring COVID-19 stress and its factors in Bangladesh: a perception-based study. Heliyon. 2020 Jul 1;6(7):p e04399.

9. Nguyen HC, Nguyen MH, Do BN, Tran CQ, Nguyen TT, Pham KM, Pham LV, Tran KV, Duong TT, Tran TV, Duong TH. People with suspected COVID-19 symptoms were more likely depressed and had lower health-related quality of life: The potential benefit of health literacy. Journal of clinical medicine. 2020 Apr;9(4):965.

10. Talevi D, Socci V, Carai M, Carnaghi G, Faleri S, Trebbi E, di Bernardo A, Capelli F, Pacitti F. Mental health outcomes of the CoViD-19 pandemic. Rivista di psichiatria. 2020 May 1;55(3):137–44.

11. Centers for Disease Control and Prevention (CDC). CDC HRQOL-14 ‘Healthy Days Measure’. Available form: https://www.cdc.gov/hrqol/hrqol14_measure.htm.

12. Directorate General of Health Services (DGHS). National Guidelines on Clinical Management of Coronavirus Disease 2019 (COVID-19), Bangladesh (version 7.0); 28 May 2020.

13. Ping W, Zheng J, Niu X, Guo C, Zhang J, Yang H, Shi Y. Evaluation of health-related quality of life using EQ-5D in China during the COVID-19 pandemic. PloS one. 2020 Jun 18;15(6):p e0234850.

14. Tran BX, Nguyen HT L.HT, Latkin CA, Pham HQ, Vu LG, L. XT, Nguyen TT, Pham QT, Ta NT, Nguyen QT. Impact of COVID-19 on economic well-being and quality of life of the Vietnamese during the national social distancing. Frontiers in psychology. 2020;11. Available form: https://dx.doi.org/10.3389%2Ffpsyg.2020.565153.

